# Participation of people with disabilities due to Leprosy, Lymphatic Filariasis (LF), and other causes in Uganda

**DOI:** 10.1101/2023.03.24.23287679

**Authors:** Carolyne Sserunkuma Maholo, Barbara Batesaki Sembatya, Kawikizi Moses, Consolata Mpambaara, Antony Sahayarani, Anil Fastenau, Srilekha Penna, Wim H. van Brakel, Christa Kasang

## Abstract

**Background:** Neglected tropical diseases (NTDs) often affect the most impoverished people in society. Uganda continues to carry a heavy burden of leprosy and lymphatic filariasis (LF). Both NTDs cause disfigurement and disability, often impacting the participation and inclusion of the affected persons. Their exclusion largely arises from the stigmatizing nature of both conditions and the limited knowledge and information. This research project conducted in 2018 investigated the participation of people with disabilities due to leprosy, LF, and other causes to inform appropriate intervention.

**Methods:** A mixed-method approach including quantitative and qualitative methods was applied. A variety of data collection methods such as Participation scale (v6.0), focus group discussions, observation, and semi-structured interviews were used to achieve comprehensive conclusions on the various aspects of participation and inclusion of people with disabilities arising from leprosy, LF, and due to other causes.

**Results:** Many people with disabilities due to leprosy and LF realize that they are affected by the disease only when it has advanced and they have developed visible disabilities. This is largely attributed to a lack of knowledge and information about the two diseases. About 27% reported no participation restriction, 25% reported severe, and 17% extreme participation restriction. Among people with disabilities due to LF a higher percentage reported no restriction, while higher percentages of respondents with disabilities due to leprosy and other causes reported severe and extreme restrictions. Participation restriction is experienced more by the female population.

**Conclusion:** Although there is a general lack of data on disability experienced by people affected by leprosy and LF, a significant percentage (42.1%) experienced participation restriction. Disability due to leprosy and other causes is more likely to cause participation restriction than disability attributed to LF. Adequate knowledge and information, accessibility, acceptability, and affordability of services are key enablers of participation while their lack hinders participation and inclusion.

**Author Summary:** Disability is estimated at 15% of an average population (WHO 2011). Disability inclusion is prioritized on the development agenda. It is explicitly mentioned in six out of the 17 goals (UN 2015). Leprosy and lymphatic filariasis are neglected tropical diseases that disfigure and affected persons and cause disability however, not much is documented on their participation in community development.

This study was implemented in 2018 to establish the nature and level of participation as well as a foundation upon which stigmatized people such as those affected by Leprosy, LF and disabilities due to other can be assured of participation and inclusion in community development programs. This mixed method study was implemented in five districts of Northern Uganda. Documents review, a participation scale, focus group discussions and semi-structured interviews were used to collect data.

This study revealed that due to lack of knowledge and information, many realize that they have leprosy and lymphatic filariasis when they have developed visible disabilities. It also revealed that people affected by leprosy, LF and PWDs access and benefit from health care services but are excluded from most mainstream community development programs. However, people affected by leprosy, people with disabilities due to other causes and the female with lymphedema reported more participation restriction than male with lymphatic filariasis. The key determinants of participation in community development are attitude, distance, funds, knowledge and information.

## Introduction and Background

Approximately 15% of the worldwide population are persons with disabilities attributed to different causes. 80% of people with disabilities live in developing countries [1]. The Uganda Bureau of Statistics Census Report indicated that 12.4% of the Ugandan population lives with some form of disability [2].

Africa is reported to account for almost 40% of the global NTD burden [3]. According to the Ministry of Health, Uganda continues to carry a heavy burden of NTDs [4]. Leprosy and Lymphatic filariasis (LF) are listed among the NTDs in Uganda. They both cause disfigurement and disability. Persons with disabilities in general are known to suffer exclusion due to limited capacity brought about by their impairment and the barriers experienced in society.

In 2018, 208 619 new cases of leprosy were reported from 127 countries compared to 211 009 cases in 2017 indicating a 1.2% global decrease [5]. According to the WHO fact sheet, 10,816 new cases were detected with grade-2 disabilities (G2D), a 1.4 per million population G2D rate [6] with visible disabilities like damage to hands and feet often used as an indicator for the severity of the leprosy disease [7] and timeliness of detection.

Leprosy is endemic in Uganda, with 40% of the districts affected in 2016 [8] and 231 new cases reported across 50 districts in 2018/19, with an annual case detection rate of 0.06/1,000,000 population. Northern Uganda accounted for over 60% of all newly notified leprosy cases [9].

In 2018, 51 million people were reported to live with LF [10]. LF impairs the lymphatic system followed by abnormal enlargements of body parts, causing pain, severe disability, and social stigma. According to the MOH 2009/2010 survey, LF was endemic in 54 districts in Uganda. Several districts in Northern Uganda were classified as needing surveillance.

This research project sought to determine the nature and extent of participation of people with disabilities attributed to leprosy, LF, and other causes and identify strategies to enhance their participation.

## Methods

### Study design

The study adopted a descriptive research design utilizing both qualitative and quantitative data. Qualitative data was used to establish the nature of participation, enablers, barriers to participation, challenges experienced, and strategies to promote participation and inclusion. Quantitative data was used to establish the various demographic aspects of the people with disabilities due to Leprosy, LF, and other causes, the magnitude of disability, and the level of participation to inform effective conclusions on the various aspects of the study.

### Study sites

The study was conducted in Kitgum, Alebtong, and Gulu (Northern Uganda) and Arua and Koboko (North Western Uganda) regions. Northern Uganda is still recovering from a 20-year civil war with the “Lord’s Resistance Army” which forced 90% of the population to abandon their homesteads for a life confined to the squalid internment camps [11]. The war increased poverty and vulnerability, leading to a general breakdown in social service provision resulting in delayed detection of NTDs and disability. On the other hand, North Western Uganda often gets an influx of refugees from Congo and South Sudan which have equally experienced war over the years and have a burden of other NTDs [12].

### Sample size and sampling

The study included people with disabilities due to leprosy who were released from treatment between 1996 and 2017 (n= 294), on treatment (n=11), and new cases (n=5). People affected by LF with different symptoms like lymphedema (n= 168) and hydrocele (n= 172), 2 people affected by leprosy and with lymphedema, and people with disabilities (PWD) due to other causes (n= 359). TB/leprosy supervisors (n=10), vector control officers (n=5), chairpersons of the disability council (n=5), chairpersons of the disabled people’s unions (n=5), community development officers (n=5), health officers (n=5), and health personnel (n=8) were included for interviews. 18 focus group discussions (FGDs) were conducted each with 15 participants including PWDs, people affected by leprosy, and LF.

While the study planned a stratified random sampling, there was a lack of reliable records of people affected by leprosy while people with disabilities and people affected by LF were never documented. As a result, scanty records of people affected by leprosy were used together with snowball sampling with the support of district TB leprosy supervisors (DTLS), village health teams, community development officers (CDOs), local leaders, and communities to identify respondents.

### Data collection

Data collection started with a document review where records from the respective health centers, NTD offices, community development offices, organizations of people with disabilities, and NTLP offices were used to identify the people affected by leprosy, LF, and people with disabilities.

Later, a questionnaire with a Participation scale was administered to the people affected by leprosy, LF, and people with disabilities to obtain statistics on demography, nature, degree of disability, need-use of assistive devices, and participation at individual, family, and community levels. Semi-structured interviews were conducted with health personnel, district community development officers, local leaders, and a few community members to identify challenges, enablers, and barriers to the participation of people affected by leprosy and LF and people with disabilities, and to determine appropriate strategies for change. FGDs were conducted in respective communities using participatory learning approaches including problem identification and analysis, stakeholder mapping, and risk and vulnerability analysis which together complemented strategy development.

### Data Analysis

Information from the Participation scale was coded, analyzed using Statistical Package for Social Science (IMB SPSS 25), and descriptively presented in response to the study objective. Information from the interviews and focus group discussions was transcribed, arranged based on emerging themes, and used to complement that from the Participation scale. The output from the document review was triangulated with data from all other data collection methods for a comprehensive understanding of the key aspects and a comprehensive conclusion of the study.

### Ethical considerations

The research proposal was assessed by Gulu University Research Ethics Committee before submission to the National Council of Science and Technology for approval (SS4994) and implementation. Informed consent was obtained from the participants and for children aged between 2-18 years and people with severe disabilities, both verbal and written consent was also obtained from their respective legal guardians.

## Results

Results from the participation scale are summarized using graphs, complemented with output from ethnography and interviews.

The majority (n=631; 62.4%) of the respondents were male with 79.8% of males in the LF group. In the interview, 63% of the respondents in the LF group indicated that they have hydrocele. Age distribution showed 132 children (7.7%), 526 (52%) respondents in the productive age (18-59 years), and 353 (34.9%) above 60 years. Further stratified distribution of the respondent’s age and gender are depicted in Figure 1.

**Figure 1:**
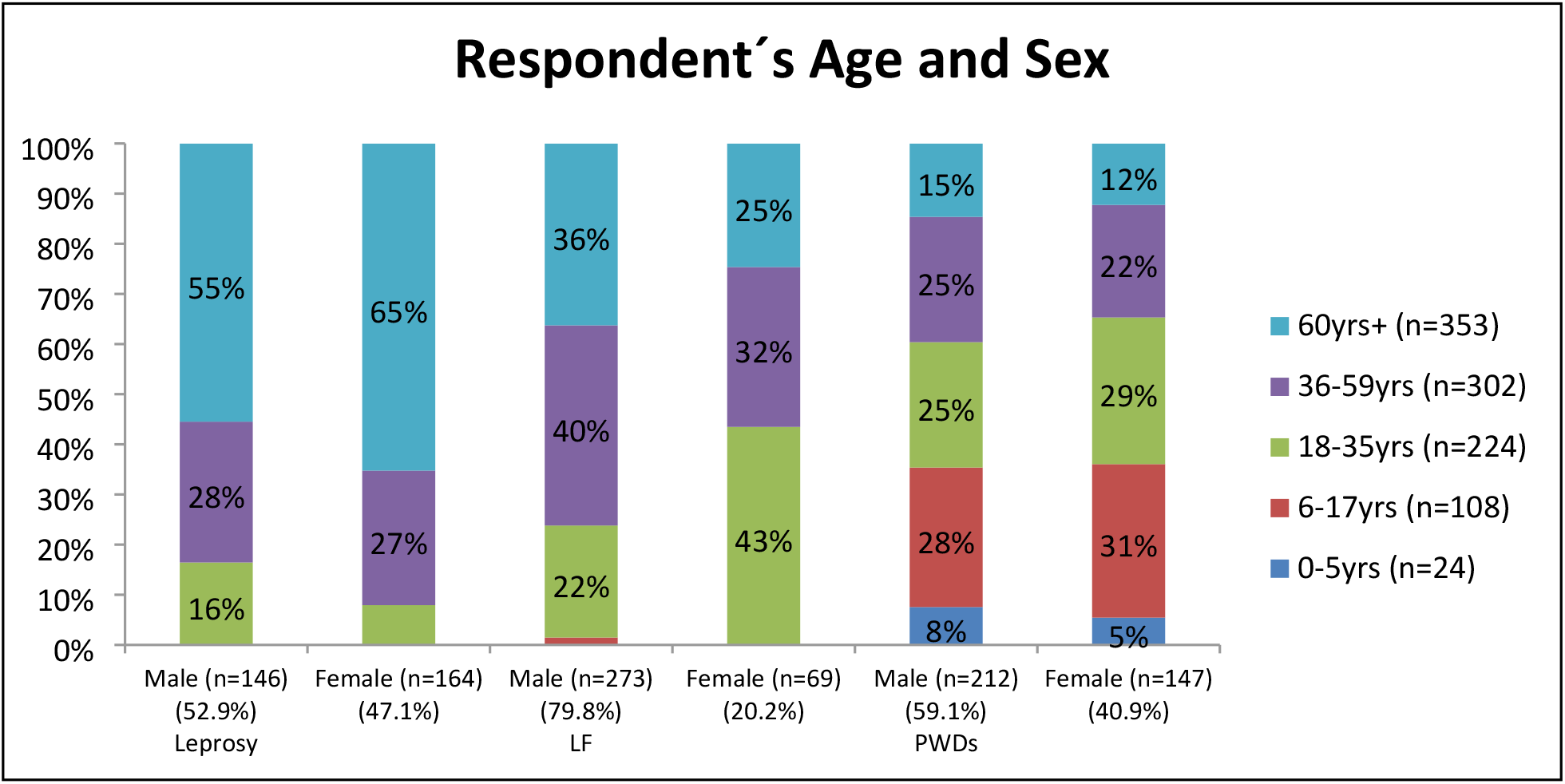
Distribution of Gender and Age among the Respondents.

To characterize the study participants best, some information on their individual perspectives of the degree of disability, use, and need of the assistive devices was obtained as detailed in Figure 2 and Figure 3.

**Figure 2:**
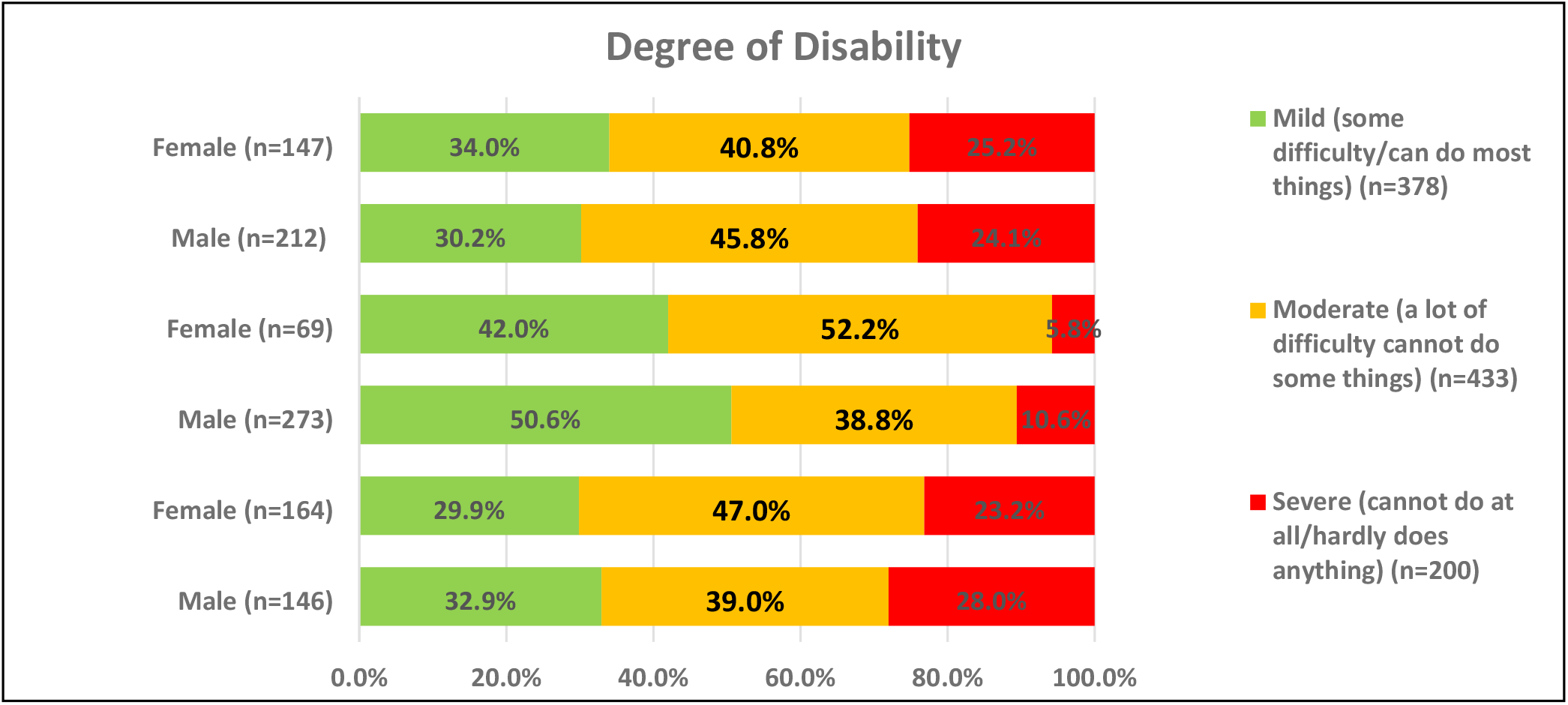
Three-category individual perspectives of the degree of disability differentiated by gender.

**Figure 3:**
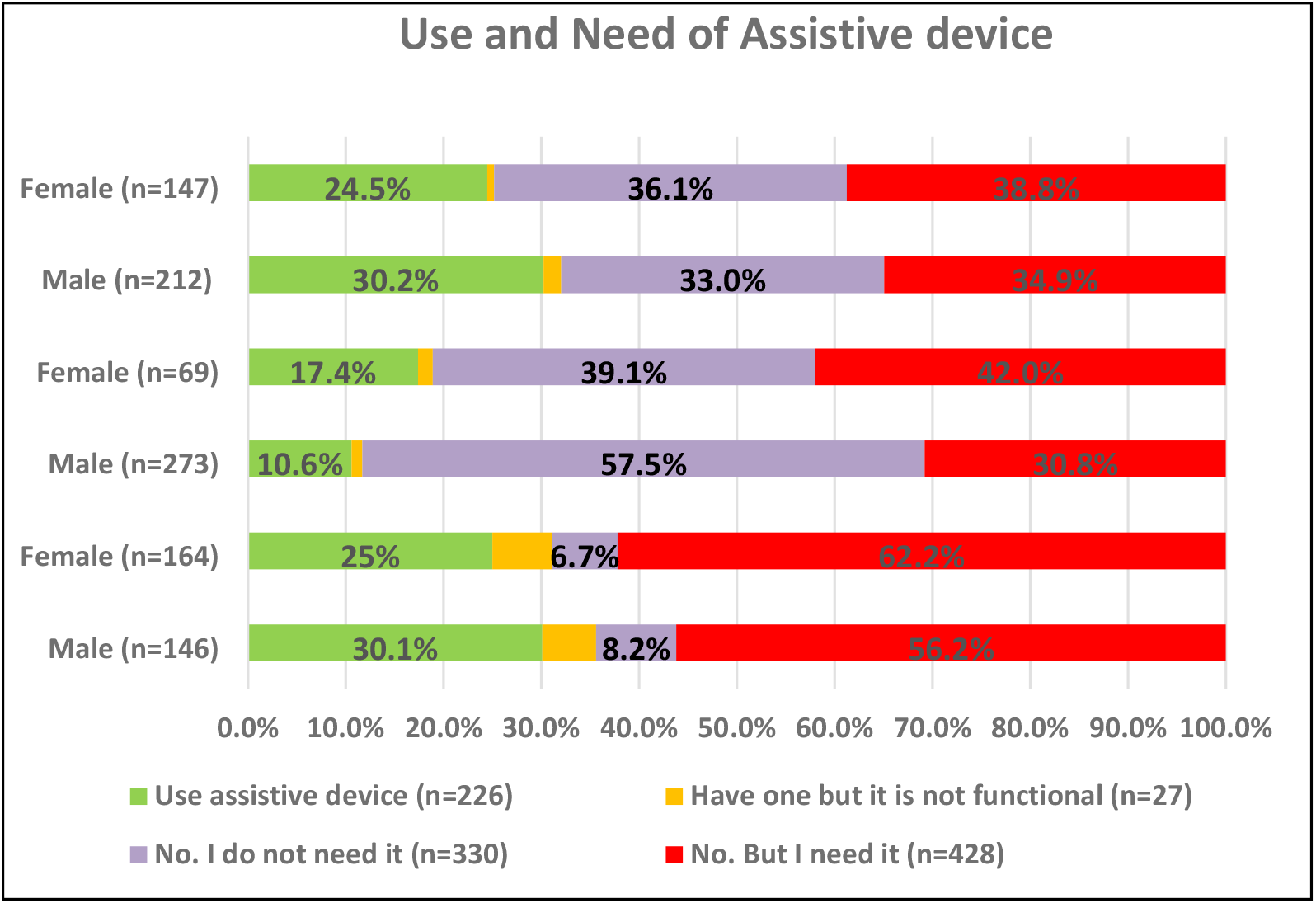
Use and need of assistive devices.

Overall 42.8% of the respondents perceived their disability to be moderate and 41.5% of the people affected by LF were in this category (Figure 2). 37.4% perceived their disability to be mild while 19.8% perceived their disability to be severe. People affected by leprosy and persons with disabilities accounted for the higher percentage of respondents with severe disabilities.

42.3% of the respondents indicated that they need assistive devices (Figure 3). Majority (59.4%) of people affected by leprosy reported the need for protective footwear and other mobility aids. Leprosy affects people’s sense of feeling and renders them vulnerable to injuries, especially in the feet and hands. The lack of assistive devices could explain the wounds commonly found with the people affected by leprosy as shown in Picture 1 and Picture 2:

**Pic1 & 2:**
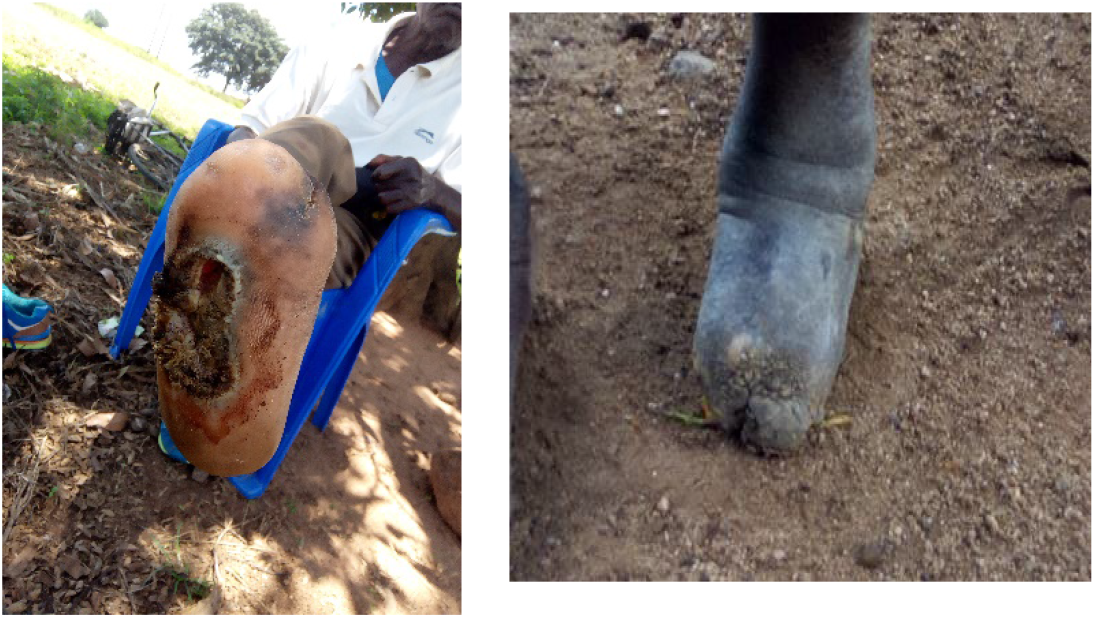
People affected by leprosy in need of protective footwear.

The person in Pic 1 was found wearing rubber sandals with a big hole at the bottom. Deep inside the septic wound was a rusty metal piece which he did not notice due to loss of sensation. The continuous wounds leave many people affected by leprosy with a bad odor which worsens their exclusion in the community.

The people with lymphedema were facing similar issues as shown in Pic 3.

**Pic3:**
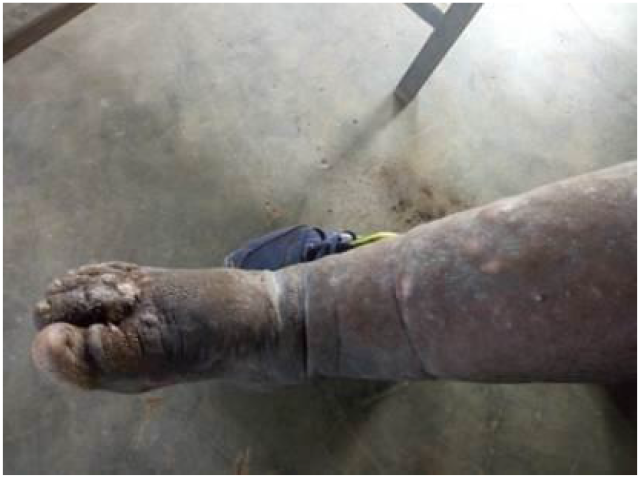
Wound of a person with Lymphedema.

Pic 3 is of a youth found during an FGD in the Gulu district. He was wearing a pair of tight jeans and canvas shoes despite the difficulty in moving. He admitted using this as a disguise for his condition to minimize exclusion as shared below:

> *“Look! I am a youth. Like all my peers, I would like to marry someday. But who can accept a man with this swollen ugly leg? I wear jeans like all youths in this village to be admired by girls. Who knows! I could get a wife soon”* (male youth with elephantiasis in Awach sub-county, Gulu district)

Participation was first assessed at the individual level, then at the family and community level using an overall participation scale (v6.0) developed by the participation development team. The results were categorized into five levels and stratified by condition and sex as shown in Figure 4.

**Figure 4:**
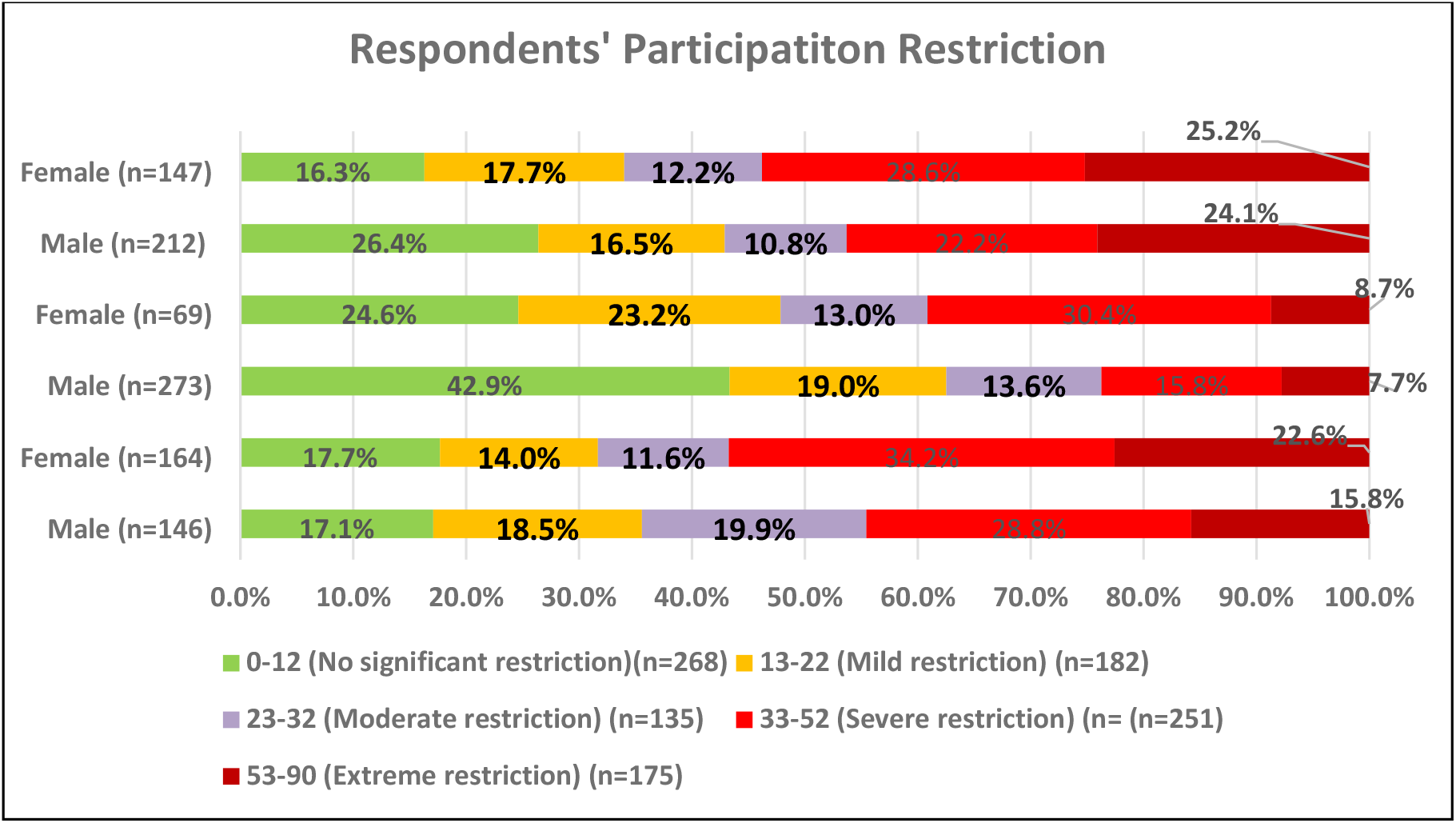
Grading of participation restriction by condition and sex.

The majority of respondents affected by leprosy (51%) and respondents with disability (49.3%) reported severe participation restrictions. In addition, a significant percentage (39.1%) of the female respondents with LF (lymphedema) reported having severe participation restrictions. However, a greater percentage of the male respondents (42.9%) with LF reported no significant participation restrictions. About 37.2% of respondents in this category perceived their disability as mild and indicated that they would not need assistive devices.

Overall, female respondents reported more participation restriction than their male counterparts. Low participation restrictions amongst males with LF could be attributed to the patriarchal nature of these communities and people’s receptive attitude to hydrocele as mentioned by a respondent below:

*‘Traditionally, a man with a hydrocele is believed to be very fertile. He can make any woman pregnant. It is a pride for the man to have that condition. It is also a sign of wealth since you make many children and girls fetch cows in this community ’* (a female respondent in an FGD in Omoro sub-county, Alebtong district)

Participation restriction was observed in all categories however, it is seen to reduce with education as shown in Figure 5. Respondents with relatively higher education (upper primary to tertiary) reported limited participation restriction. 36.5 % of the respondents have never been to school and 86.6% of the female affected by leprosy were in this category. Of these, 51.8% were older persons of 60 and above who most likely suffered from leprosy in the early years worsened by the exclusion of women at the time.

**Figure 5:**
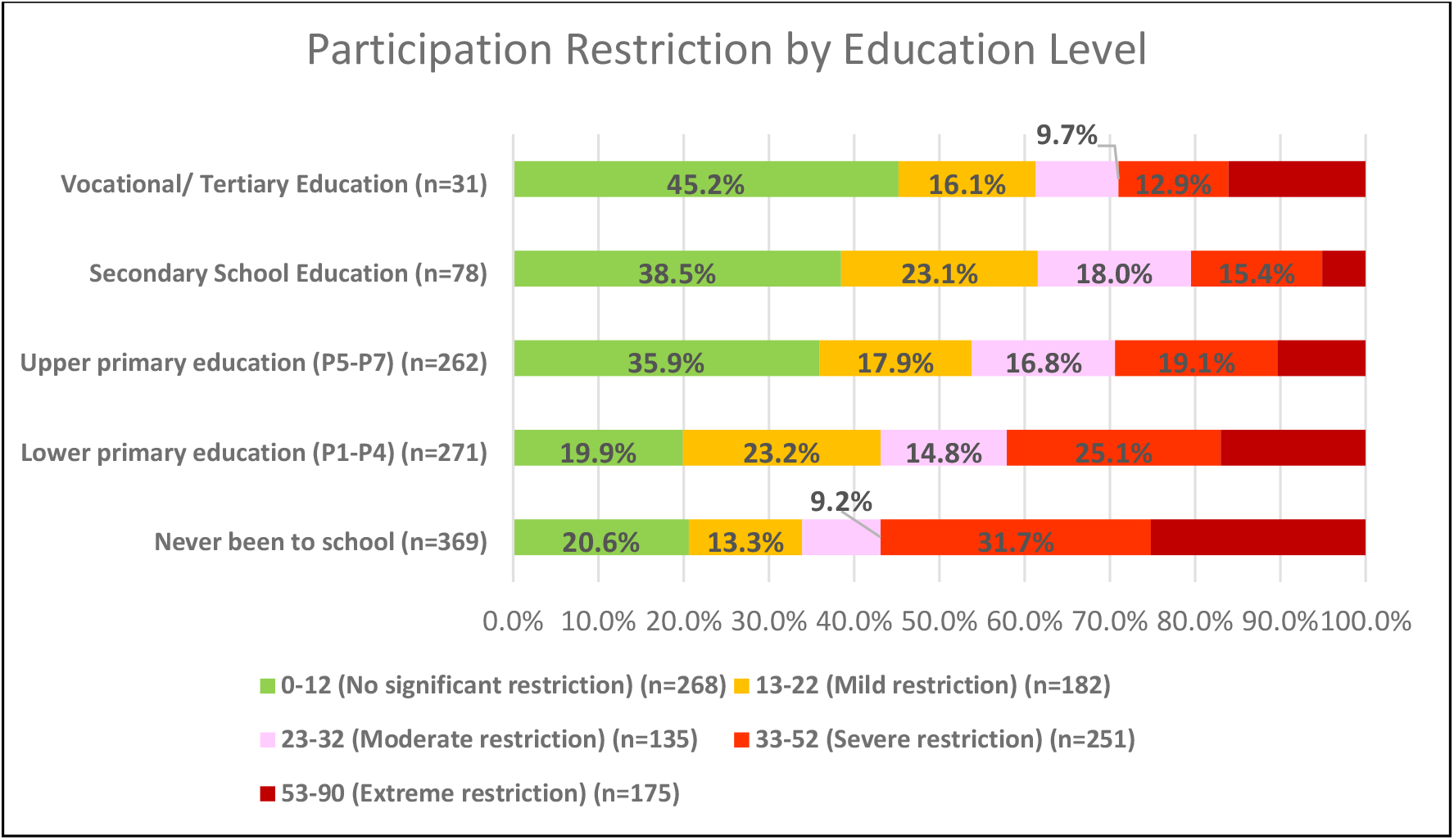
Grading of participation by the level of education.

Appropriate intervention requires a detailed exploration of the various areas in which individual categories experience participation restriction. All the above responses were based on individual perspectives, attitude, and readiness to participate. Figure 6 depicts the details of the participation of respondents in different daily life situations.

**Figure 6:**
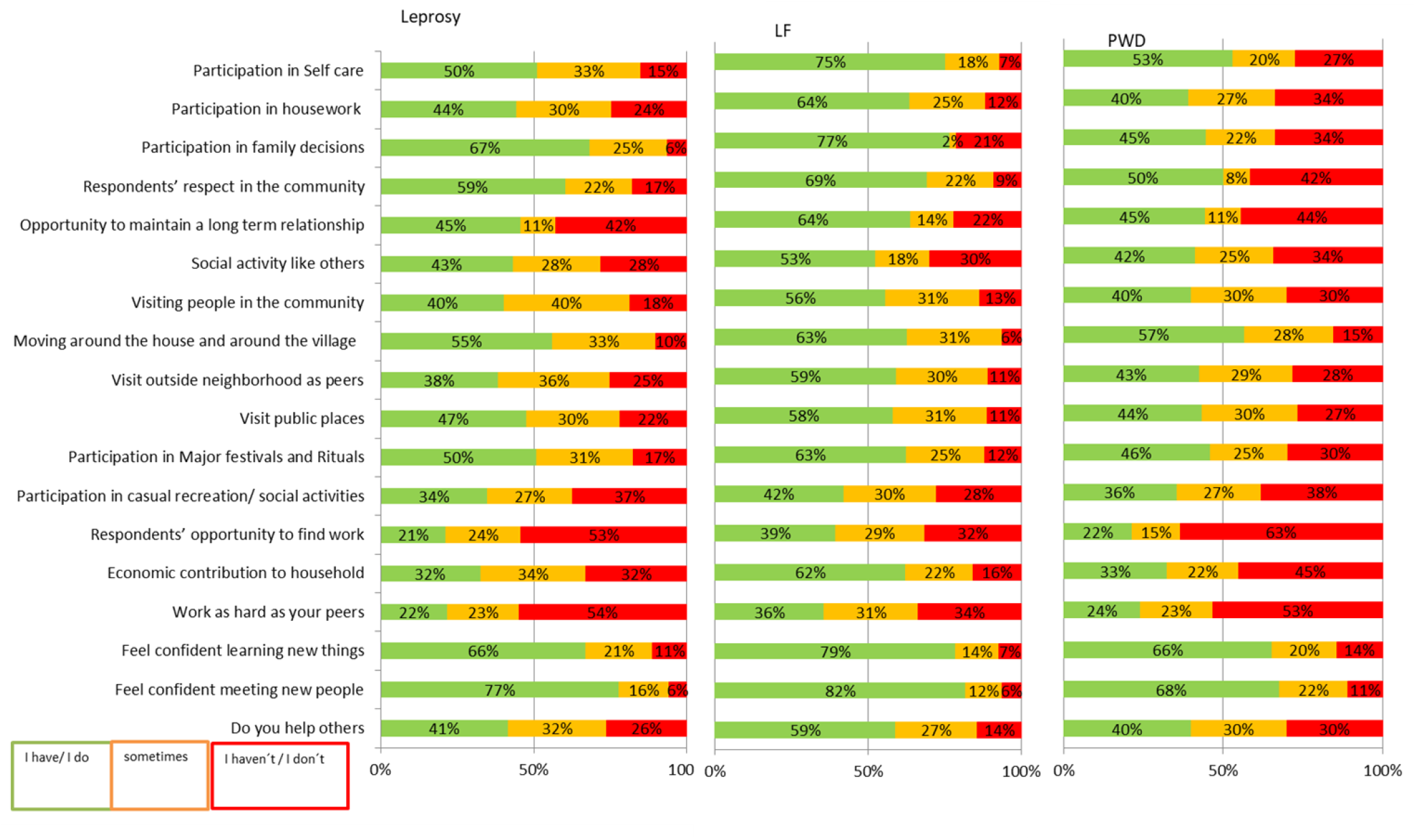
Participation in different daily life situations.

74.7% of respondents reported that they are comfortable meeting new people, 69.1% are confident to try learning new things, 65.1% mentioned their opinions counted in the family, 60.4% care for themselves, and 58.5% are respected in the community. Most of these were reported by male respondents.

Participation restriction was reported in finding work (44.2%), working as hard as others (41.8%), participating in casual recreation activities, and being socially active. The majority of people affected by leprosy (54%), persons with disabilities (63%), and people with lymphedema (60.4%) reported restrictions on finding work, which could be an affirmation of exclusion in economic participation.

Generally, the people affected by leprosy, persons with disabilities, and people with lymphedema who participated in the study indicated more participation restriction than those with hydrocele. This could be attributed to the receptive attitudes that some communities have towards hydrocele since it is associated with wealth and high fertility as shared by one of the youths in the Alebtong district:

> *“In this community, this is a treasure (pointing at his scrotal area) why operate it when I can make any woman pregnant? Who doesn’t treasure children?”* (youth with hydrocele in Abako sub-county, Alebtong district)

67% of the people affected by leprosy reported participation in decision making and 59% have respect in the community. Since the majority (60.7%) of the people affected by leprosy were older persons of 60 years and above, this could be attributed to the respect of older persons as custodians of indigenous knowledge.

60.4% of the respondents reported self-care. Although this was reported in all three categories of respondents, people affected by leprosy and people with lymphedema had poor hygiene. They would easily be spotted during the FGDs because of the bad odor and attraction of houseflies. In one group, a lady affected by leprosy wore stockings but not even her peers (affected by leprosy) wanted her near them because of the odor she came with.

> *“Let her sit there alone!” the group unanimously shouted*.

> *“How can a normal woman be that smelly? You are the reason we get rejected in the community. Why don’t you clean up and wash those wounds? We all have them but no…*..*you will have to sit there alone”*

The lady reported that she wears stockings to prevent flies on the wounds on her feet. It also helps her hide her condition.

Further, strategies to enhance the participation of the people affected by leprosy, LF, and persons with disabilities were obtained by triangulating information from all data collection methods. Results showed that many people only realize they have leprosy when it has already advanced and they have disabilities as shared below:

> *“Patients usually come to the health facility when leprosy has advanced and with deformity. In such cases, we treat leprosy but the disability is left for life unless intervention comes in to address it. Raising awareness of leprosy will inform people so that they report to health facilities early. It will help to reduce new cases and reduce disabilities due to leprosy”* (medical personnel in Koboko district)

During the FGDs, people affected by leprosy and LF mentioned the need for training on self-care at individual and group levels as a means of mitigating the bad odor that most people had. Having realized the similarities that leprosy and LF have to a disability, the disability councilors, disability union officials, and community development officers agreed to promote the inclusion of people affected by leprosy and LF in community development programs.

## Discussion

Our study revealed that people affected by leprosy and LF experience activity limitation [13], and are vulnerable and excluded as people with disabilities due to other causes [14]. About half (51%) of the people affected by leprosy and a high percentage (49%) of the respondents with disabilities reported severe participation restriction. Likewise, a substantial percentage (39%) of the female respondents with lymphedema reported severe participation restriction [15] while most of their male counterparts experienced no significant participation restriction. This shows that disability and the disfigurement caused by leprosy and lymphedema restrict the social and work participation of the affected persons. Much as the exclusion of people affected by leprosy could be attributed to unfavorable attitudes, limited knowledge, and information, it could also be due to poor access to assistive devices as 59% of persons affected who participated in the study indicated a lack of these.

Half of the respondents who reported extreme participation restriction and severe restrictions have never been to school. According to the most recent National Population census, Northern Uganda where the study was undertaken registered high percentages of individuals who did not complete primary education [16]. The majority (60%) of the female respondents who had never been to the school indicated severe and extremely severe participation restrictions, which could imply double vulnerability and exclusion attributed to gender and disability as a result of these three conditions. 87% of the women affected by leprosy were in this category and of these, 52% were older persons (60 and above). The 2016/2017 Uganda household survey reported that 42% of older persons have never been to school and 57% of these are female [17]. Lack of education among older women could have resulted from the exclusion of women before the United Nations Decade for Women (1975 to 1985) which gave rise to policies and strategies to promote, equity, stop gendered violence and all their rights [18] which worsened by the myths associated with leprosy at the time [19]. Education is a great empowering tool since it enlightens individuals, builds confidence, and breaks barriers to opportunity [20]. Thus, our study revealed that education is one of the key determinants of participation at different levels.

Half of the people affected by leprosy, the majority (63%) of the people with disabilities, and people with lymphedema (60.4%) reported restriction in finding work which could be an affirmation of exclusion in economic participation. Since this rating was based on individual perceptions, participation restriction could be attributed to intrinsic and extrinsic attitudes due to the stigmatizing nature of these three conditions. A similar study reported that participation restrictions and stigma, shame, problems related to marriage, and difficulties in employment were the most frequently reported problems experienced by people disabled by leprosy [21].

However, 67% of the people affected by leprosy mentioned that they participate in decision making and 59% said they have respect in the community which could be attributed to age since 61% of the people affected by leprosy were older persons (60 years and above). In many communities of Uganda, older persons are respected as custodians of indigenous knowledge and contribute to the caregiving of orphans and other vulnerable children [22].

60.4% of the respondents reported that they participate in self-care. Although this was observed in all three categories of respondents, most of the people affected by leprosy and people with lymphedema had poor hygiene and sanitation which contributed to their exclusion. There is a definite need to strengthen self-care skills among those affected by leprosy and LF.

The provision of knowledge and information was identified to help with the early identification and referral of the affected persons for appropriate intervention. In a similar study, knowledge, and information on leprosy were reported to be instrumental in behavior change communication to facilitate attitude change [23]. Overall, training in self-care, inclusion in disability programs, and awareness creation on leprosy, LF, and disability were also identified as priorities to promote participation and inclusion of people affected by leprosy, LF, and persons with disabilities.

## Conclusion

People affected by leprosy and LF experience as much activity limitation, vulnerability, and exclusion as persons with disabilities due to other causes. People with hydrocele have no significant participation restriction while the people affected by leprosy and people with lymphedema experience as much participation restriction as other persons with disabilities.

Knowledge and information, attitude, and self-care appear to be important determinants of participation and inclusion of people affected by leprosy, LF, and disability due to other causes, while lack of reliable documentation restricts effective planning for their inclusion.

## Recommendations

Uganda published disability-inclusive planning guidelines [24] to ensure disability inclusion in all government programs. However, its successful adoption requires effective documentation of people affected by leprosy, LF, and persons with disabilities, which is currently lacking.

Therefore, proper documentation of the people affected by leprosy, LF, and persons with disabilities is necessary to inform effective planning and intervention to ensure their participation and inclusion. This can start by use of the lists generated by this study.

Furthermore, the adoption of Community Based Rehabilitation (CBR) with a focus on self-care is essential to empower individuals affected by leprosy, LF, and persons with disabilities. The multisectoral collaboration and use of local resources in CBR will contribute to the mitigation of barriers to ensure their inclusion.

People affected by leprosy, LF, and persons with disabilities should be mobilized into groups to benefit from the existing disability and mainstream programs since this is the country’s strategy to promote inclusive development. This can be boosted with the enhancement of capacity for awareness creation, management of injuries and related challenges, self-advocacy, collective saving and investment, and livelihood development for empowerment and enablement.

To mitigate leprosy and disability due to leprosy, there is a need to intensify capacity building in the identification, assessment, management, and referral of suspected cases of leprosy. To bridge the knowledge gap, health education, and health camps should also be organized using community-based approaches to demystify leprosy, LF, and disability. Similar studies should be replicated in all the other districts of the country.

Generally, it is vital to engage the disability community to recognize disability that arises from leprosy, LF, and other neglected tropical diseases for collective advocacy to promote participation and disability inclusion in contribution to the sustainable development goals.

## Data Availability

Data will be accessed from the Kyambogo University Institutional Repository

## Acknowledgments

The Research Team expresses sincere gratitude to the Leprosy Research Initiative for the opportunity, technical and financial support to successfully implement this study. Appreciation goes to German Leprosy and TB Relief Association for the coordination of this study; UNALEP team for the support throughout data collection, enlightening communities on leprosy and contributing to the fight against leprosy; the District Health Officers, District TB Leprosy Supervisors, NTD personnel, Community Development Officers, the leadership in the disability unions and disability councils, the health personnel, members of the village health teams and local leaders who greatly committed time to support the study.

Special gratitude goes to the people affected by leprosy, Lymphatic Filariasis and persons with disabilities, their families and the community for the time committed and the willingness to share the details presented in this report. Mr Okello Edwin and Mr Faraday M. Michael Faraday for the tremendous skills, determination and resilience throughout data management to facilitate successful completion of the assignment.

Finally, we also acknowledge all colleagues who have helped to review draft offering useful advice.

## Author Contributorship

Conceptualization: KM, BBS; CK research concept and methodology, MCS, CK; data collection MCS, BBS, KM; data management and analysis MCS, CK; report writing MCS; validation and technical guidance WHV, CK; logistics and administration CM; review of report and writing article AS, CK, AF, SP and WHV.

## Supporting Information (if any)

## Additional Information

## Financial Disclosure Statement

This research project was implemented with financial support from the Leprosy Research Initiative *with grant number 706*.*18*.*46*.

The funders had no role in study design, data collection and analysis, decision to publish, or preparation of the manuscript.

## Competing Interests

All the authors declare that there is no competing interest.

## Related Manuscripts

All the authors certify that the article is their original work and has not been submitted for publication or review in any other journal.

## Notes

### Competing Interest Statement

The authors have declared no competing interest.

### Author Declarations

The research proposal was assessed by Gulu University Research Ethics Committee before submission to the National Council of Science and Technology for approval (SS4994) and implementation.

